# Antibody responses boosted in seropositive healthcare workers after single dose of SARS-CoV-2 mRNA vaccine

**DOI:** 10.1101/2021.02.03.21251078

**Authors:** Todd Bradley, Elin Grundberg, CODIEFY study team, Rangaraj Selvarangan

**Author notes:** Co-Corresponding Authors: Rangaraj Selvarangan, Todd Bradley. CODIEFY study team (in alphabetical order): Dithi Banerjee, Bradley Beldon, Rebecca Biswell, Elizabeth Fraley, Cas LeMaster, Daniel Louiselle, Angela Myers, Nick Nolte, Tomi Pastinen, Jennifer Schuster.

## Abstract

Current guidelines recommend that individuals who have had COVID-19 should receive the identical vaccine regimen as those who have not had the infection. This includes two doses of the mRNA platform vaccines (BNT162b2/Pfizer; mRNA-1273/Moderna) that are approved for use in the United States. In this brief report, we show that after a single dose of the Pfizer SARS-CoV-2 vaccine, individuals that had prior SARS-CoV-2 infection had significantly higher antibody levels than individuals that had no history of infection. This provides the rationale for changing vaccination policy to deliver only a single dose to individuals with recent SARS-CoV-2 infection that may free up additional doses for individuals that have no preexisting immunity to the virus. Future study of other immune parameters such as T cell response and durability of immune response should be rapidly undertaken in individuals that had COVID-19 prior to vaccination.

## Main

Currently, there are two SARS-CoV-2 vaccines approved for emergency use by the FDA that utilize mRNA platform technology (BNT162b2/Pfizer; mRNA-1273/Moderna) ^1,2^. Phase 3 trials of these vaccines showed greater than 90% efficacy at preventing symptomatic infection after two doses administered three to four weeks apart. These trials were conducted primarily in participants without previous SARS-CoV-2 infection. There have been over 26 million documented cases of COVID-19 in the US with high rates of seroprevalence observed in recent studies^3^. Thus, there is an urgent need to determine the immune response to vaccination in persons with previous SARS-CoV-2 infection. Due to current shortages in global vaccine production and distribution, single dose regimens in certain populations, such as those with preexisting immunity, may increase vaccine availability.

Here we determined antibody levels at baseline and 3 weeks after the first vaccine dose of the Pfizer BNT162b2 SARS-CoV-2 mRNA vaccine in healthcare workers with laboratory confirmed SARS-CoV-2 infection 30-60 days prior to vaccine (COVID19+; N=36) or workers without history of infection (COVID19-;N=152). Using a multiplex bead binding assay (Millipore #HC19SERG1-85K) that measures IgG levels against SARS-CoV-2 spike protein subunits S1, S2, Receptor Binding Domain (RBD) and the nucleocapsid protein (NP), we found that after the 1^st^ vaccine dose, both COVID19- and COVID19+ individuals antibody titers were enhanced to all proteins with the exception of NP which is not a vaccine antigen (P<0.0001, Wilcoxon-Mann-Whitney; Figure 1A). At baseline, we observed 6 individuals that had antibody levels that matched COVID-19 seropositive status and may have had undiagnosed infection. Namely, having MFI ≥1000 for the S1 protein. We separated these individuals from analysis (Undiagnosed) and found that they resembled the COVID19+ in the week 3 serology assays. After the first vaccine dose COVID19+ individuals had significantly higher antibody titers to the S1, S2 and RBD compared to COVID19-individuals (P<0.0001, Wilcoxon-Mann-Whitney). As a proxy for measuring virus neutralizing antibodies, we utilized an FDA approved *in vitro* assay that allows qualitative detection of SARS-CoV-2 neutralizing antibodies in the blood (Genscript, #L00847; Figure 1B). We found that neutralizing antibodies were boosted for the COVID19+ individuals (median 48.6% blocking at baseline and median 96.3% blocking at week3; P<0.0001, Wilcoxon-Mann-Whitney). While COVID19-individuals neutralizing antibodies were also boosted after vaccination (median 8.9% blocking at baseline and median 59.5% blocking at week3) they were significantly lower than the COVID19+ individuals (P<0.0001, Wilcoxon-Mann-Whitney).

**Figure 1.**
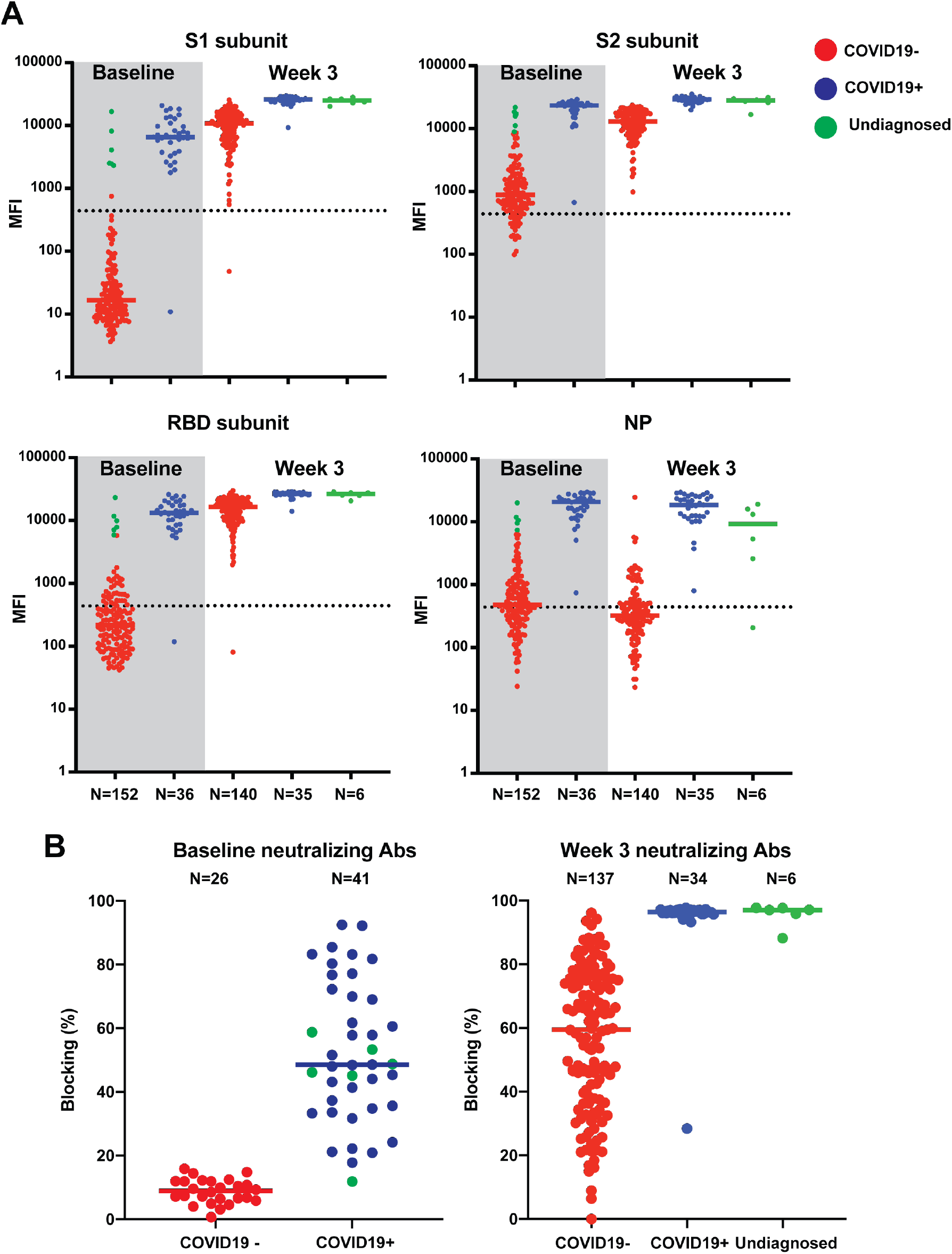
Antibody response to SARS-CoV-2 mRNA vaccine. **(A)** Multiplex bead-based antibody binding assay that measures the IgG antibody response to 4 SARS-CoV-2 viral antigens (S1, S2, RBD and NP). Each dot represents an individual at baseline before vaccine or 3 weeks after the first dose of vaccine. Bars represent group median. Number of individuals in each group are shown below the graphs. Individuals with history of infection (blue), no history of infection (red) and possible undiagnosed infection (green). Dashed line indicates limit of detection determined by the sum of the average and standard deviation of the negative control (beads without antigen). **(B)** Neutralization antibody proxy assay that determines the level of antibodies that block the RBD-ACE2 receptor binding. Each dot represents an individual at baseline before vaccine or 3 weeks after the first dose of vaccine. Bars represent group median. Number of individuals in each group are shown below the graphs. Individuals with history of infection (blue), no history of infection (red) and possible undiagnosed infection (green). Black circles at baseline indicate Undiagnosed individuals.

These findings suggest that in individuals with recent SARS-CoV-2 infection or seropositive status elicit a more rapid booster antibody response to vaccination and provides a rationale for considering a single dose vaccine regimen in this population. The duration of antibody responses also will need further investigation.

## Data Availability

Primary data are available from the authors at reasonable request.

## Ethics statement

The vaccinee biospecimens were collected under a clinical study at Children’s Mercy Kansas City and reviewed and approved by the Children’s Mercy IRB (#00001670 and #00001317).

## Acknowledgements

We thank all the health care workers that participated in this study. Special thanks to Occupational Health and CM Research Institute for their support of this study.

## References

1. Baden LR, El Sahly HM, Essink B, et al. Efficacy and Safety of the mRNA-1273 SARS-CoV-2 Vaccine. N Engl J Med 2020.

2. Polack FP, Thomas SJ, Kitchin N, et al. Safety and Efficacy of the BNT162b2 mRNA Covid-19 Vaccine. N Engl J Med 2020;383:2603–15.

3. Lumley SF, O’Donnell D, Stoesser NE, et al. Antibody Status and Incidence of SARS-CoV-2 Infection in Health Care Workers. N Engl J Med 2020.

